# Incidence, Risk Factors and Mortality Outcome in Patients with Acute Kidney Injury in COVID-19: A Single-Center Observational Study

**DOI:** 10.1101/2020.06.24.20138230

**Authors:** Alfano Gaetano, Ferrari Annachiara, Fontana Francesco, Mori Giacomo, Magistroni Riccardo, Meschiari Marianna, Franceschini Erica, Menozzi Marianna, Cuomo Gianluca, Orlando Gabriella, Santoro Antonella, Di Gaetano Margherita, Puzzolante Cinzia, Carli Federica, Bedini Andrea, Milic Jovana, Raggi Paolo, Girardis Massimo, Mussini Cristina, Cappelli Gianni, Guaraldi Giovanni, for the Modena Covid-19 Working Group (MoCo19)

**Affiliations:** Surgical, Medical and Dental Department of Morphological Sciences, Section of Nephrology, University of Modena and Reggio Emilia, Modena, Italy; Nephrology Dialysis and Transplant Unit, University Hospital of Modena, Modena, Italy; Clinical and Experimental Medicine PhD Program, University of Modena and Reggio Emilia, Modena, Italy; Clinic of Infectious Diseases, University Hospital of Modena, Modena, Italy; Department of Surgical, Medical, Dental and Morphological Sciences; Department of Medicine, Division of Cardiology, Mazankowski Alberta Heart Institute, Alberta Canada; Department of Anesthesia and Intensive Care Unit, Azienda Ospedaliero-Universitaria Policlinico of Modena, Modena, Italy; Infectious Diseases Clinics, University Hospital, via del Pozzo 71, 41124 Modena, Italy; Department of Anesthesia and Intensive Care, University Hospital, via del Pozzo 71, 41124 Modena, Italy; Chair of Pathology and Immunology, University of Modena and Reggio Emilia, Via Campi, 287, 41125 Modena, Italy

## Abstract

**Background:** Acute kidney injury (AKI) is a recently recognized complication of coronavirus disease-2019 (COVID-19). This study aims to evaluate the incidence, risk factors and case-fatality rate of AKI in patients with documented COVID-19.

**Methods:** We reviewed the health medical records of 307 consecutive patients hospitalized for symptoms of COVID-19 at the University Hospital of Modena, Italy.

**Results:** AKI was diagnosed in 69 out of 307 (22.4%) patients. The stages of AKI were stage 1 in 57.9%, stage 2 in 24.6% and stage 3 in 17.3%. Hemodialysis was performed in 7.2% of the subjects. AKI patients had a mean age of 74.7±9.9 years and higher serum levels of the main marker of inflammation and organ involvement (lung, liver, hearth and liver) than non-AKI patients. AKI events were more frequent in subjects with severe lung comprise. Two peaks of AKI events coincided with in-hospital admission and death of the patients. Kidney injury was associate with a higher rate of urinary abnormalities including proteinuria (0.448±0.85 vs 0.18±0.29; P=<0.0001) and hematuria (P=0.032) compared to non-AKI patients. At the end of follow-up, 65.2% of the patients did not recover their renal function after AKI. Risk factors for kidney injury were age, male sex, CKD and non-renal SOFA. Adjusted Cox regression analysis revealed that AKI was independently associated with in-hospital death (hazard ratio [HR]=3.74; CI 95%, 1.34-10.46) compared to non-AKI patients. Groups of patients with AKI stage 2-3 and failure to recover kidney function were associated with the highest risk of in-hospital mortality. Lastly, long-hospitalization was positively associated with a decrease of serum creatinine, likely due to muscle depletion occurred with prolonged bed rest.

**Conclusions:** AKI was a dire consequence of patients with COVID-19. Identification of patients at high-risk for AKI and prevention of kidney injury by avoiding dehydration and nephrotoxic agents is imperative in this vulnerable cohort of patients.

## Introduction

COVID-19 is a complex infectious disease characterized by a broad spectrum of manifestations ranging from asymptomatic to severe illness^1^. The disease is associated with a high rate of morbidity and mortality in patients hospitalized for severe symptoms of SARS-CoV-2 pneumonia^2^. Patients older than 65 years and people of all ages with a high burden of comorbidities have the highest risk of severe complications and poor outcome^3–5^. Lung is the main target of the virus, but other organs including brain, liver and kidneys can be involved in this infection^6^. To date, the pathogenesis of COVID-19 is poorly understood and the principal etiology of organ dysfunction seems due to the direct and indirect effects of the proinflammatory cytokine^7–9^.

The rate of kidney injury in COVID-19 is unclear, but recent evidence has established that kidney involvement is parallel to the severity of the underlying lung involvement^10^. Therefore, the estimated prevalence of acute kidney injury (AKI) may be proportional to the severity of the host systemic immune response. Studies conducted in China and the US have reported a high prevalence of urinary abnormalities (proteinuria and microhematuria) and a rate of AKI ranging from 0.5 to 36.6%^10–16^ of the patients. A recent report from Bordeaux (France) documented that the impact of acute renal injury has been estimated to about 80% in severely ill patients admitted in ICU^17^.

The etiological mechanisms leading to kidney injury are still unknown. There are three potential interlinked mechanisms: direct cytopathologic damage, cytokine storm and drug toxicity. Kidney biopsies performed on 26 patients with COVID-19 who died of multiple organ failure showed widespread proximal tubule injury consistent with acute tubular necrosis^18^. The detection of the virus in urine leads to hypothesize a potential cytopathic effect of the virus within kidneys, but the direct demonstration of the virus in renal parenchyma remains an area of open debate requiring future investigations. In addition, there are no data on the effect of immune-modulating drugs on kidney damage in SARS-CoV-2.

Understanding the impact of SARS-CoV-2 on renal function is necessary to elucidate the epidemiological and clinical characteristics and the survival of patients experiencing AKI. The aim of this study was to evaluate the incidence, risk factors of AKI in COVID-19 and its association with mortality.

## Methods

### Study design and setting

This retrospective, observational study was conducted in patients with laboratory confirmed-COVID-19 admitted to the University Hospital of Modena. The city of Modena is geographically located in Emilia Romagna region that overall accounted for a total amount of 28.143 documented COVID-19 cases as of 18 June 2020^19^. Clinical and laboratory data were prospectively recorded in consecutively admitted patients from 23 February to 27 April 2020.

The study was approved by the regional ethical committee of Emilia Romagna (prot. n. 0013376/20)

### Population

This study recruited all consecutive adult patients (≥18 years) admitted with SARS-CoV-2 infection. Patients with ESRD in renal replacement therapy were excluded from the analysis. According to the WHO guidelines, the diagnosis of SARS-CoV-2 was defined as a positive real-time reverse transcriptase-polymerase chain reaction (RT-PCR) assay of nasopharyngeal swabs or lower respiratory tract specimens. ^20^

At the end of the follow-up, the patients were classified as alive, dismissed, or dead.

### Standard of care

Delivery of healthcare services for all SARS-CoV-2 infected patients was ensured by a public healthcare system. Care of COVID-19 patients was delivered by an integrated multidisciplinary team including infectious disease specialists, pneumologists, internal medicine physicians, nephrologists, rheumatologists, and intensive care, and coagulation specialists.

According to the Italian Society of Infectious Diseases’ Guidelines (SIMIT) ^21^ and recent data on the treatment of COVID-19 ^22,23^ all patients received standard of care treatment including: (a) oxygen supply to achieve SaO2 ≥ 90%; (b)hydroxychloroquine (400 mg BID on day 1 followed by 200 mg BID on days 2 to 5); (c) azithromycin (500 mg QD for 5 days); (d) Darunavir/cobicistat (800/150 mg QD) for 14 days; (e) low molecular weight heparin for prophylaxis of deep vein thrombosis.

From 18 March 2020 combined therapy darunavir/cobicistat was stopped due to the supervening information on the lack of clinical benefit of protease inhibitors to treat COVID-19, lopinavir in particular^24^. A sub-cohort of patients received tocilizumab treatment in addition to the standard of care when they met the following criteria: SO_2_<92% and a PaO_2_/FiO_2_<200 mmHg in room air or a decrease in PaO2/FiO2 > 30% in the previous 24 hours after hospitalization.

Severely ill patients were evaluated by intensive care consultants for ICU admission and invasive mechanical ventilation eligibility. Medical history, age, comorbidities, vital signs, physical and laboratory examinations were assessed daily.

### Criteria and definition

AKI was defined according to the 2012 Kidney Disease: Improving Global Outcomes (KDIGO) criteria^25^. Three AKI stages were classified as follow:

(i) stage 1: increase in serum creatinine (sCr) ≥0.3 mg/dl within 48 hours or 1.5 to 1.9 times increase of baseline sCr measured within 7 days; (ii) stage 2: 2-2.9 times increase of baseline sCr measured within 7 days; (iii) stage 3: 3 times or greater increase in baseline sCr measured within 7 days or initiation of RRT. ^25^ AKI stage was the highest reached during hospitalization. Urine output criteria were not used consistently in all patients for the diagnosis of AKI due to a lack of daily urine measurement in the electronic chart.

Baseline sCr was defined as the last available sCr measurement within 365 days before the onset of COVID-19 symptoms. When not available prior to the diagnosis of COVID-19, sCr measured on admission was used as the ‘baseline’ value.

The estimated glomerular filtration rate (eGFR) was calculated using the Chronic Kidney Disease Epidemiology Collaboration (CKD-EPI) equation^26^.

Polypharmacy was considered present when five or more medications were used^27^.

Non-renal SOFA was calculated by subtracting the score resulting from the degree of renal dysfunction from total score^28^.

Requirements for admission to the ICU were: (i) hypoxia despite noninvasive ventilatory support; (ii) hemodynamic instability; (iii) cardiac arrest; (iv) respiratory arrest; (v) multiorgan failure.

### Data collection

Data collected from electronic medical records included demographics, comorbidities, medications administered at home and in hospital, laboratory values, vital signs and outcomes. They were prospectively recorded from hospital admission. Comorbidities were identified upon review of the patient’s medical records. International Classification of Diseases (ICD) was used to code and classify mortality data from death certificates.

### Outcome

The primary outcome measure was the incidence of AKI in hospitalized patients with COVID-19. Additional analyses included the detection of risk factors for AKI and its relationship with mortality among hospitalized patients.

### Statistical analysis

Baseline characteristics were analyzed using descriptive statistics and were reported as proportions or mean (standard deviation [SD]). χ2 test or Fisher’s test was used to analyze categorical variables. Analyses of continuous variables were compared using an unpaired t-test or Kruskal-Wallis test, as appropriate. A gamma distribution function was used to plot the probability of AKI events during hospitalization. Correlation and linear regression were performed to verify the relationship between the decrease of sCr and days of hospitalization in non-AKI patients. In this group of patients, the decrease of sCr was the difference between last sCr and baseline creatinine; analysis were performed only with values of last sCr equal to or less than baseline sCr.

Mortality and incidence of AKI were calculated using Kaplan-Meier (K-M) curves. Univariate and multivariate analysis was performed by Cox regression to identify risk factors for AKI. Cox regression identified independent predictors of all-cause mortality, adjusted for sex, age at diagnosis, CKD, CVD, diabetes and COPD.

A P value of less than 0.05 was considered statistically significant. SPSS 23®was used for statistical analysis.

## Results

### Clinical characteristic of patients with AKI

A total of 307 patients were included in the study. During the study period, 22.4% (n =69) of patients developed AKI. The mean age of patients with AKI was 74.7±9.9 years. sCr was measured 734 times in the AKI group (10.6 times per patient) during the average period of hospitalization lasting 16.7±10.6 days.

Mean baseline sCr in AKI group was 1.08±0.5 mg/dl, peaking 2.6±1.8 mg/dl after 9.3±7.9 days from admission. Patients with AKI reported a higher rate of proteinuria (0.448±0.85 vs 0.18±0.29; P=<0.0001) and hematuria (P=0.032) compared to patients without AKI. The main markers of organ involvement (BNP, troponin, AST, INR) and systemic inflammatory response (IL-6, c-reactive protein [CRP], ferritin) were significantly higher than non-AKI group (Table 1 and 2). The rate of mechanical ventilation and ICU admission in AKI group were 26 and 34.7%, respectively. Overall, patients with severe acute respiratory distress syndrome requiring mechanical ventilation had a higher rate of AKI events than non-mechanically ventilated patients (P=0.045). In particular, the incidence of AKI stage 2 and 3, and unrecovered AKI was higher in patients on mechanical ventilation (Supplementary Table 1)

**Table 1.** Demographics and lab examinations of AKI and non-AKI patients.

| Variable | All patients*<br>(n=307) | AKI*<br>(n=69) | No-AKI*<br>(n = 238) | AKI vs. no-AKI*<br>P-value |
| --- | --- | --- | --- | --- |
| Age, years | 65.2 ± (14.01) | 74.7± 9.9 | 62.4±13.8 | **< 0.0001** |
| Range | 25-94.4 | 43-94.2 | 25-94.4 |  |
| Males, n. (%) | 219 (71.3) | 55 (79.7) | 164 (68.9) | 0.096 |
| Race/ethnicity |  |  |  |  |
| White | 298 (97) | 68 (98.5) | 230 (96.6) | 0.689 |
| Black | 7 (2.2) | / | 7 (2.9) | 0.35 |
| Other | 2 (0.6) | 1(1.4) | 1 (0.4) | 0.399 |
| Lab test, mean (±SD) |  |  |  |  |
| Hemoglobin, g/l | 11,9 | 11.2±2.1 | 12.2±1.76 | **< 0.001** |
| White cells, mm3 | 9175±3650 | 11454±7805 | 8160±4304 | 0.542 |
| Platelets, 10<sup>9</sup>/l | 264.3±128.7 | 231.7±122.2 | 276.2±129 | **< 0.0001** |
| Glycemia | 106±47.8 | 111.7±47.5 | 104.1±47.7 | 0.14 |
| Potassium, mmol/l | 4.6±3 | 4.01±0.6 | 4.01±3.3 | 0.991 |
| Sodium, mmol/l | 137.6±4 | 139±5.52 | 137.2±3.5 | **< 0.001** |
| Calcium, mg/l | 8.5±0.5 | 8.4±0.578 | 8.6±0.5 | 0.873 |
| Albumin, gr/dl | 2.9±0.5 | 2.7±0.49 | 3.06±0.47 | **< 0.0001** |
| Urea, mg/dl | 59±41.7 | 87.8±55.3 | 45.9±24.4 | **<0.0001** |
| D-dimer, mg/l | 6064.2±7183.6 | 5090.2±6588 | 3130±4969 | **< 0.0001** |
| Alanine amino-transferase, U/l | 136.7±827.1 | 339±161 | 67.8±78.8 | **< 0.0001** |
| Bilirubin, mg/l | 0.8±0.7 | 0.99±1.2 | 0.74±0.58 | **< 0.001** |
| INR | 1.15±0.4 | 1.33±0.73 | 1.09±0.16 | **< 0.0001** |
| Lactate dehydrogenase, U/l | 960.8±3173.1 | 1782.4±6183 | 679.2±350.7 | **< 0.0001** |
| Ferritin ng/ml | 944.8±684.3 | 958.6±691.7 | 914.3±683.3 | 0.824 |
| CPK, U/l | 211.1±687.8 | 285.2±1071 | 187.6±507.4 | **0.008** |
| BNP, pg/ml | 129.3±194.6 | 221.6±273 | 98.4±148.3 | **< 0.0001** |
| Troponin, ng/ml | 173.3±1239 | 276.6±1775 | 77.01±183.7 | 0.338 |
| IL-6, ng/L | 531.1±684.6 | 784.8±888 | 444.4±575.3 | **< 0.0001** |
| C-reactive protein, mg/l | 6.78±7.8 | 6.87±7.9 | 6.74±7.8 | 0.737 |
| Kidney function |  |  |  |  |
| sCr measurements | 2542 | 734 | 1808 |  |
| Number of sCr/pt | 8,2 | 10,6 | 7,6 |  |
| Baseline sCr, mg/dl | 0.97±0.58 | 1.08±0.5 | 0.8±0.2 | **<0.0001** |
| Range | 0.24-3.78 | 0.46-3.78 | 0.24-2.19 |  |
| eGFR, ml/min | 83.6±22.3 | 69.3 ±21.8 | 87.75±20.8 | **<0.0001** |
| Range | 14.8-147 | 14.8-111.9 | 28.5-147 |  |
| Peak sCr, mg/dl | 1.3±1.1 | 2.6±1.8 | 0.9±0.2 | **<0.0001** |
| Nadir sCr, mg/dl | 0.7±0.5 | 1.1±0.8 | 0.6±0.2 | **<0.0001** |
| Urine protein-to-creatinine ratio | 0.27±0.57 | 0.44±0.88 | 0.18±0.29 | **<0.0001** |
| Spot urine Na<sup>+</sup> | 97.93±58.48 | 75.11±40.69 | 80.27±39.64 | 0.763 |
| Spot urine K<sup>+</sup> | 36.97±20.57 | 40±15.04 | 42.24±27.52 | 0.813 |
| Hematuria |  |  |  |  |
| Absent | 99(32.2) | 12 (26.6) | 87 (36.5) | **0.032** |
| ±/+ | 31 (10) | 19 (42.2) | 22 (16) | **<0.0001** |
| ++ | 14 (4.5) | 4 (8.8) | 10 (7.2) | 0.525 |
| +++ | 11 (3.5) | 2 (4.4) | 9 (6.5) | 0.098 |
| ++++ | 17 (5.5) | 8 (17.7) | 9 (6.5) | **0.03** |

AKI, acute kidney injury; BNP, Brain Natriuretic Peptide; CKD, chronic kidney disease; CPK, creatine phosphokinase; eGFR, estimated glomerular filtration rate; IL-6, interleukin-6; INR, international normalized ratio; sCr, serum creatinine.
\* Results are expressed as mean±standard deviation, unless differently indicated.

**Table 2.**
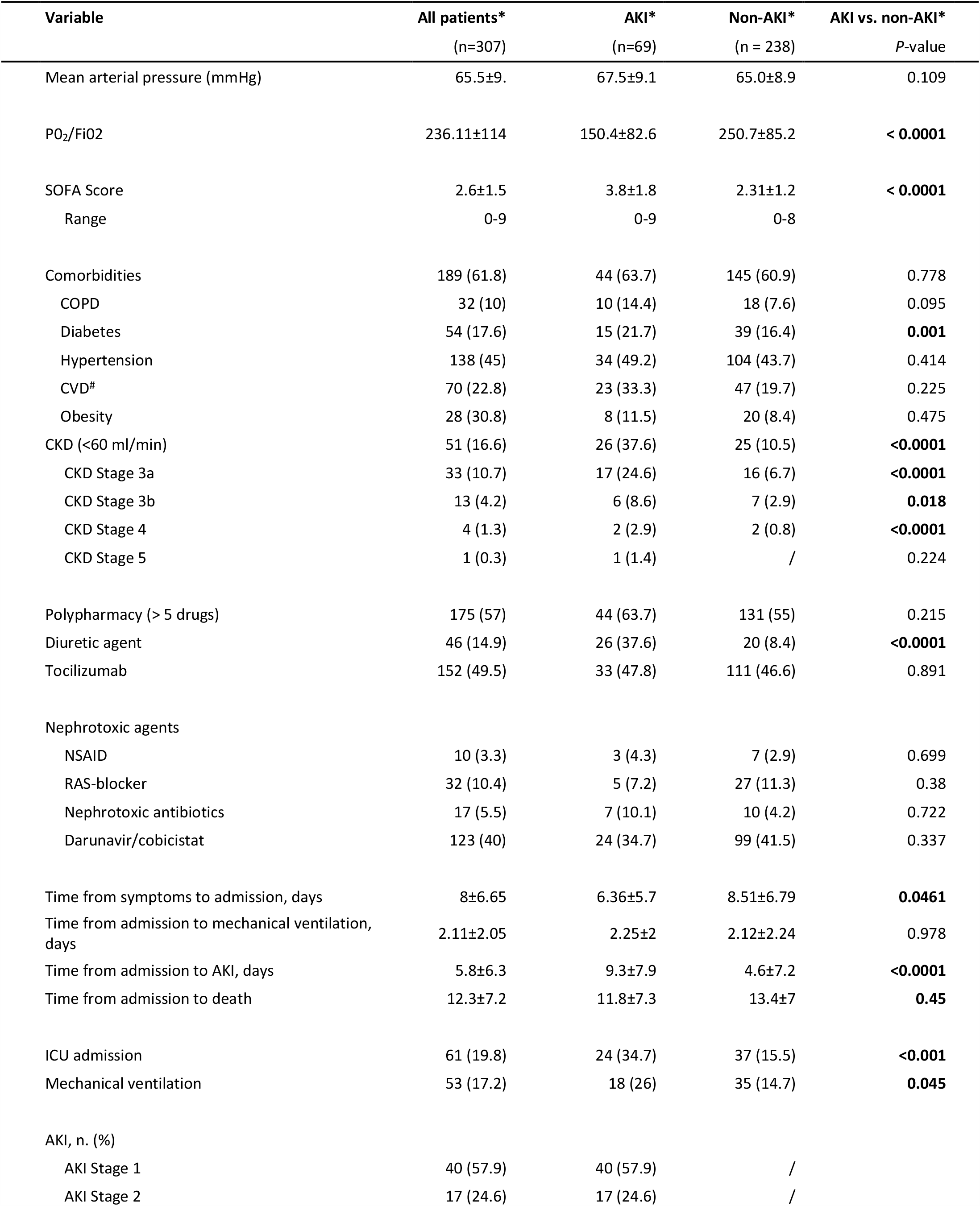
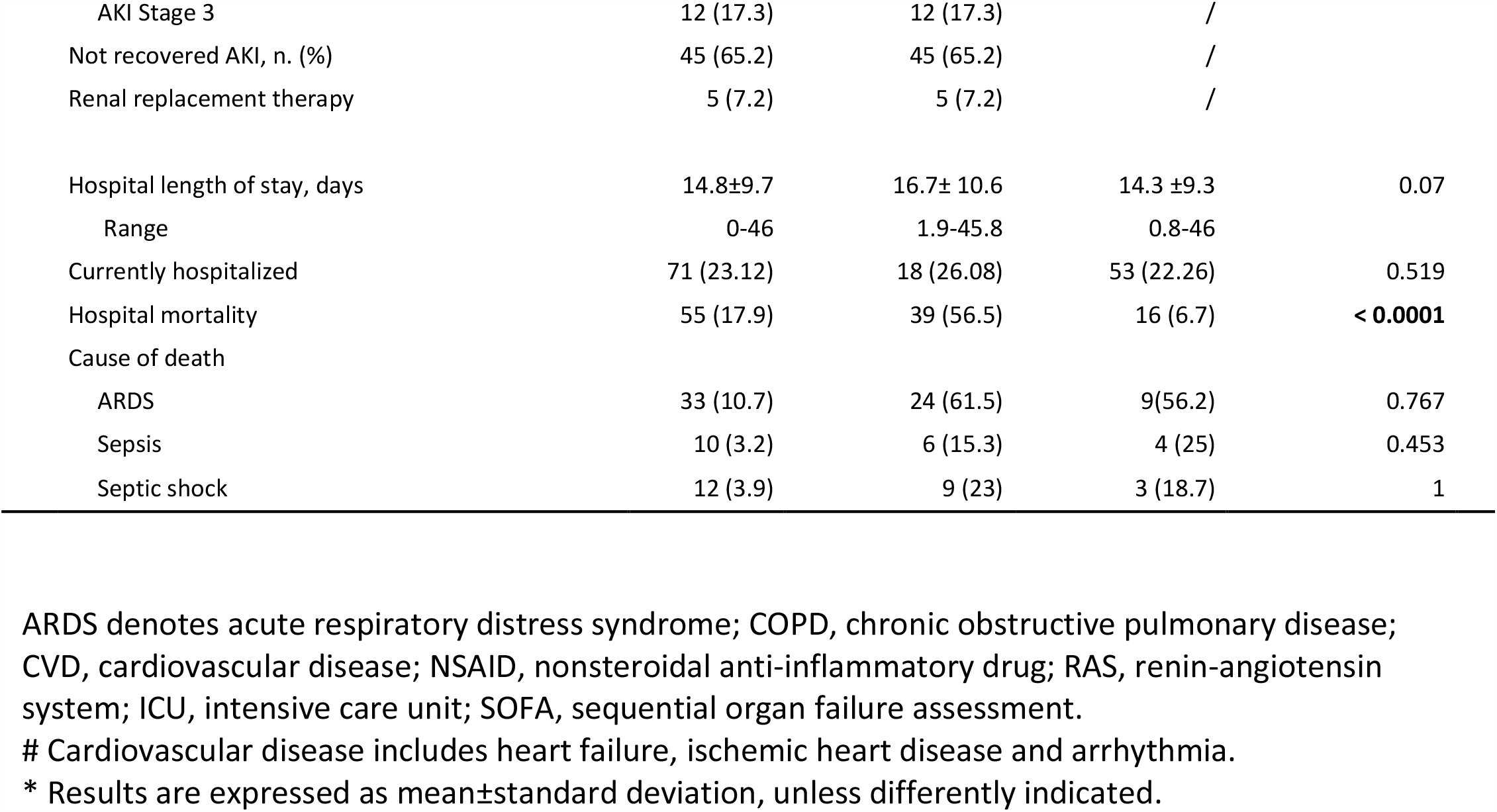
Clinical characteristic and clinical outcome of AKI and non-AKI patients.

**Table 3.** Univariate Cox Regression analysis to identify predictors of AKI.

| Variable | HR | CI (95%) |  | P-value |
| --- | --- | --- | --- | --- |
| Sex |  |  |  |  |
| Male | 1.64 | 0.91 | 2.95 | 0.100 |
| Age (1-yr increase) | 1.06 | 1.04 | 1.08 | **<0.0001** |
| ≥65 years | 4.24 | 2.23 | 8.09 | **<0.0001** |
| <65 years | 0.24 | 0.12 | 0.45 | **<0.0001** |
| Comorbidity | 1.09 | 0.67 | 1.78 | 0.727 |
| Hypertension | 1.10 | 0.69 | 1.77 | 0.683 |
| Diabetes | 1.24 | 0.70 | 2.19 | 0.470 |
| CVD | 1.76 | 1.06 | 2.90 | 0.028 |
| COPD | 1.57 | 0.80 | 3.08 | 0.188 |
| Obesity | 1.52 | 0.63 | 3.68 | 0.352 |
| CKD | 2.88 | 1.76 | 4.71 | **<0.0001** |
| GFR< 45ml/min | 2.39 | 1.19 | 4.83 | 0.015 |
| GFR 45-59 ml/min | 2.28 | 1.31 | 3.97 | 0.004 |
| Nephrotoxic antibiotic | 0.95 | 0.43 | 2.12 | 0.903 |
| RAS-blocker | 0.62 | 0.25 | 1.53 | 0.296 |
| NSAID | 1.11 | 0.35 | 3.54 | 0.860 |
| Darunavir/Cobicistat | 0.97 | 0.58 | 1.63 | 0.914 |
| Polypharmacy | 1.01 | 0.59 | 1.73 | 0.974 |
| Tocilizumab | 0.77 | 0.48 | 1.24 | 0.278 |
| No-renal SOFA (1-point increase) | 1.61 | 1.36 | 1.92 | **<0.0001** |
| Score ≤ 3 | 0.20 | 0.09 | 0.42 | **<0.0001** |
| Score > 3 | 5.10 | 2.41 | 10.80 | **<0.0001** |

CKD denotes chronic kidney disease; COPD, chronic obstructive pulmonary disease; CVD, cardiovascular disease; NSAID, nonsteroidal anti-inflammatory drug; RAS, renin-angiotensin system; SOFA, sequential organ failure assessment

### Stage of AKI

Patients with AKI were stratified according to the 2012 KDIGO guidelines and were distributed as follows: stage 1: 57.9%, stage 2: 24.6%, and stage 3: 17.3%. At the end of follow-up 65.2% of the patients did not return their initial renal function as prior to admission. Five patients with AKI stage III progressed to dialysis (chronic venous-venous hemodialysis [CVVH] or hemodialysis [HD]) in the intensive care unit (ICU); all five died as a consequence of multi-organ failure (MOF). Figure 1 shows the cumulative incidence and the distribution of AKI events during the course of the disease. The cumulative incidence curves show a steep rise in events within the first 10-15 days from admission. The graphic depiction of all AKI events shows two peaks in the timeline distribution of cases: an early peak is around the time of admission and a late one around 13-15 days from the start of hospital care (Fig. 2).

**Figure 1.**
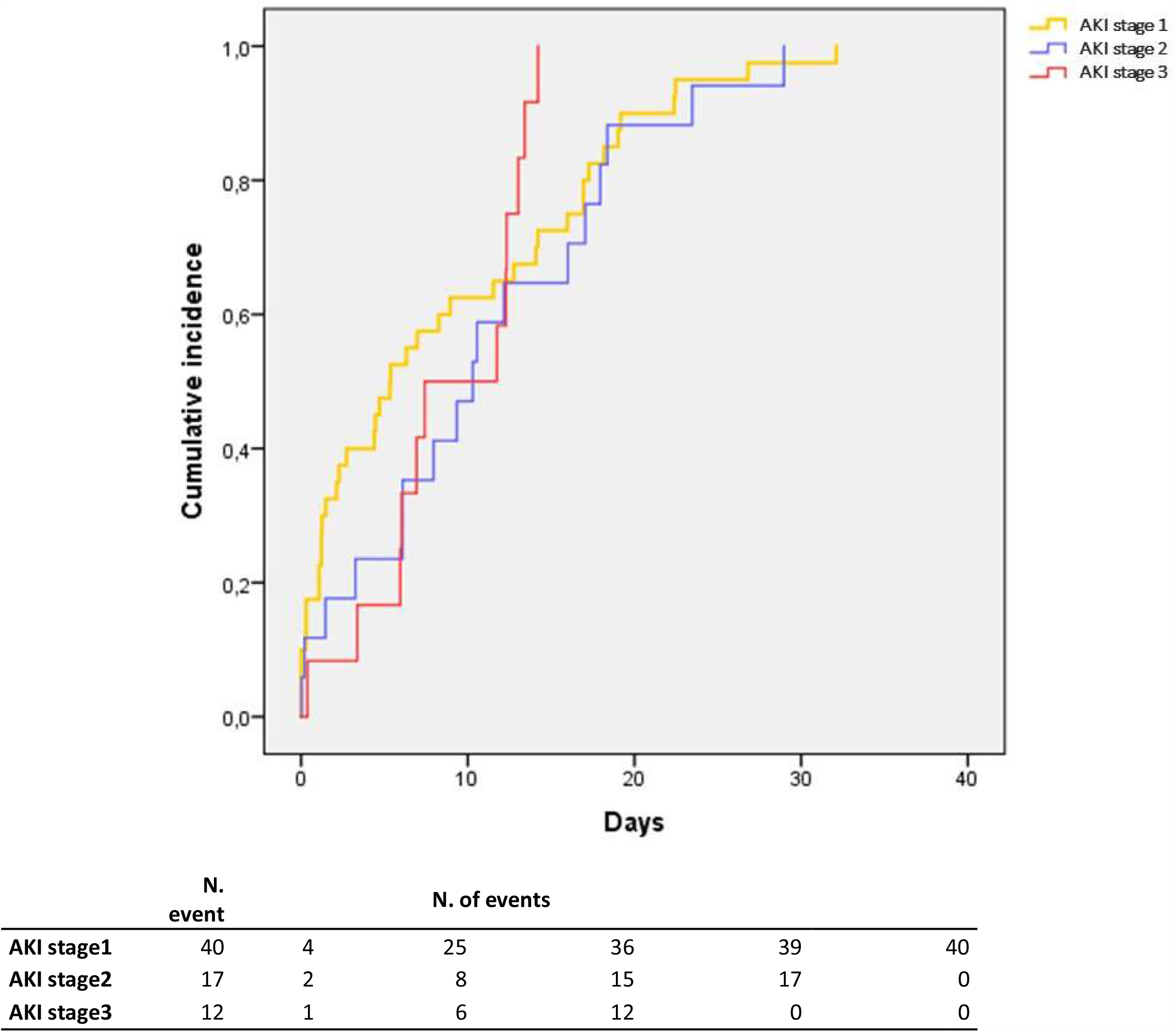
Cumulative incidence of AKI stratified according to KDIGO stage 1-3 during hospitalization.

**Figure 2.**
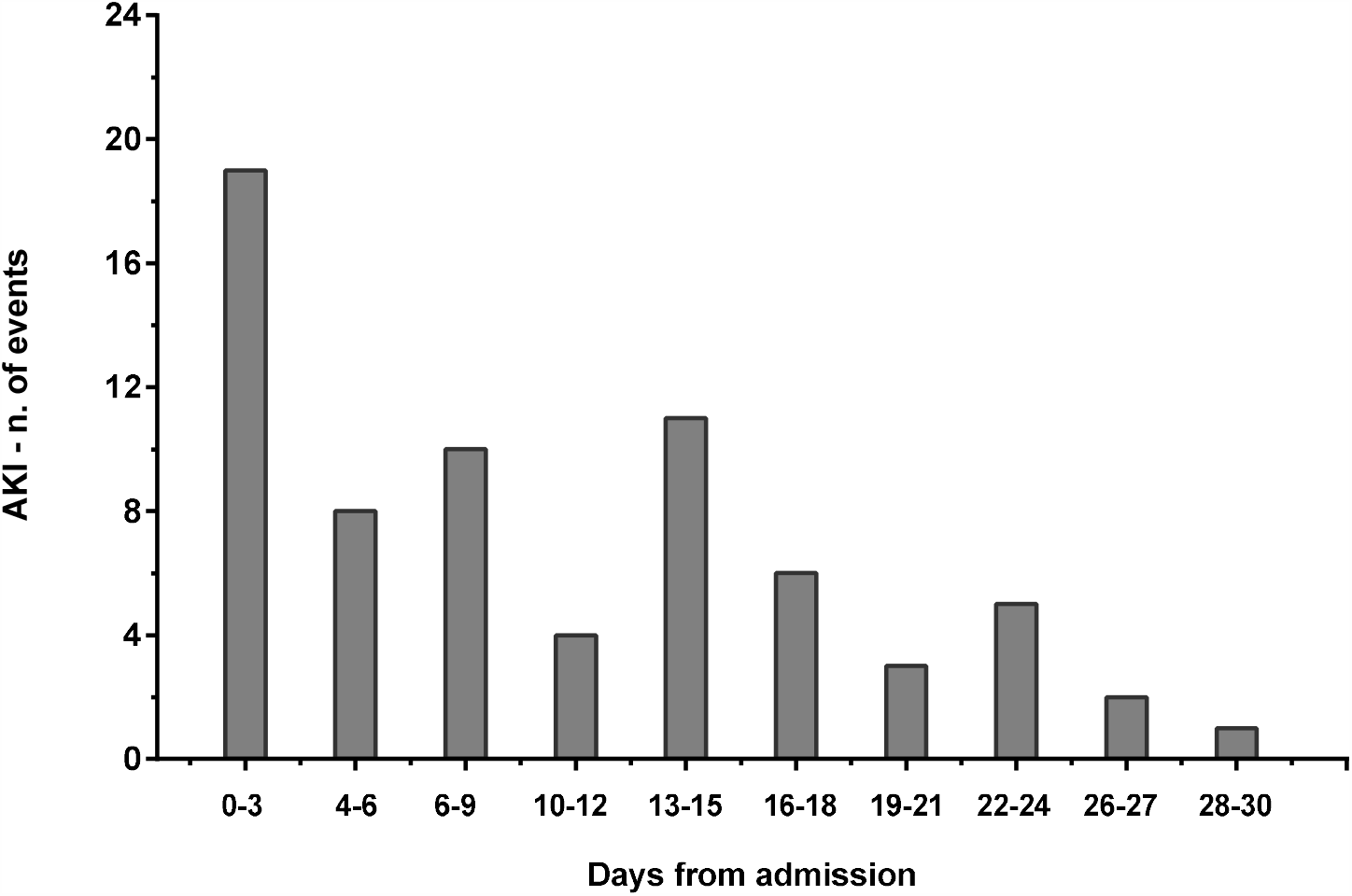
Incidence of AKI events during hospitalization for COVID-19.

Observation of the probability distribution plot (Fig. 3) revealed that the probability of developing AKI coincided with the timing of hospital admission and death of the patients occurring on average 11.8 days after admission.

**Figure 3.**
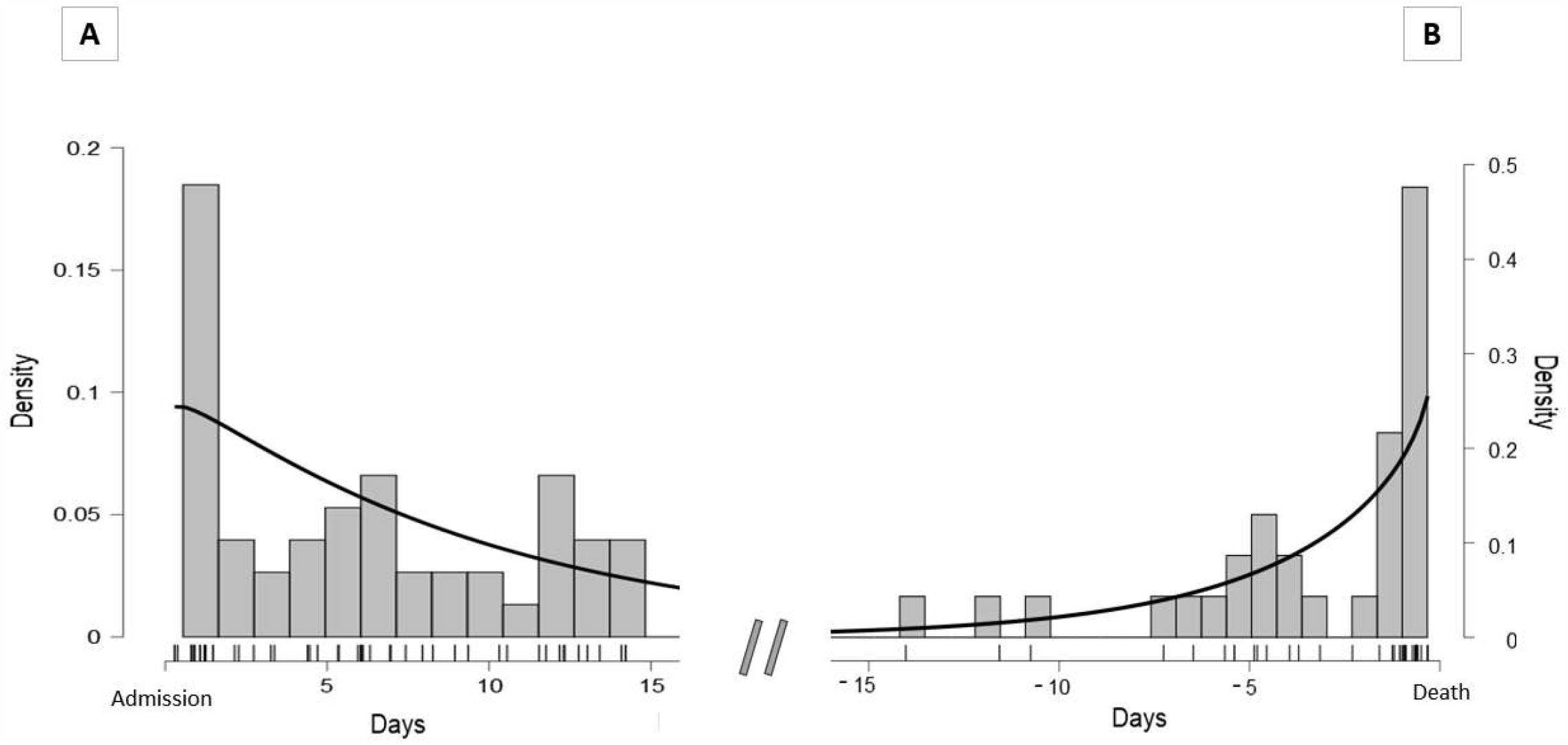
Probability of AKI related to the hospital admission (A) and death (B). In the Figure 2B, the plot of the probability density function has been performed only on non-survivals.

**Figure 4.**
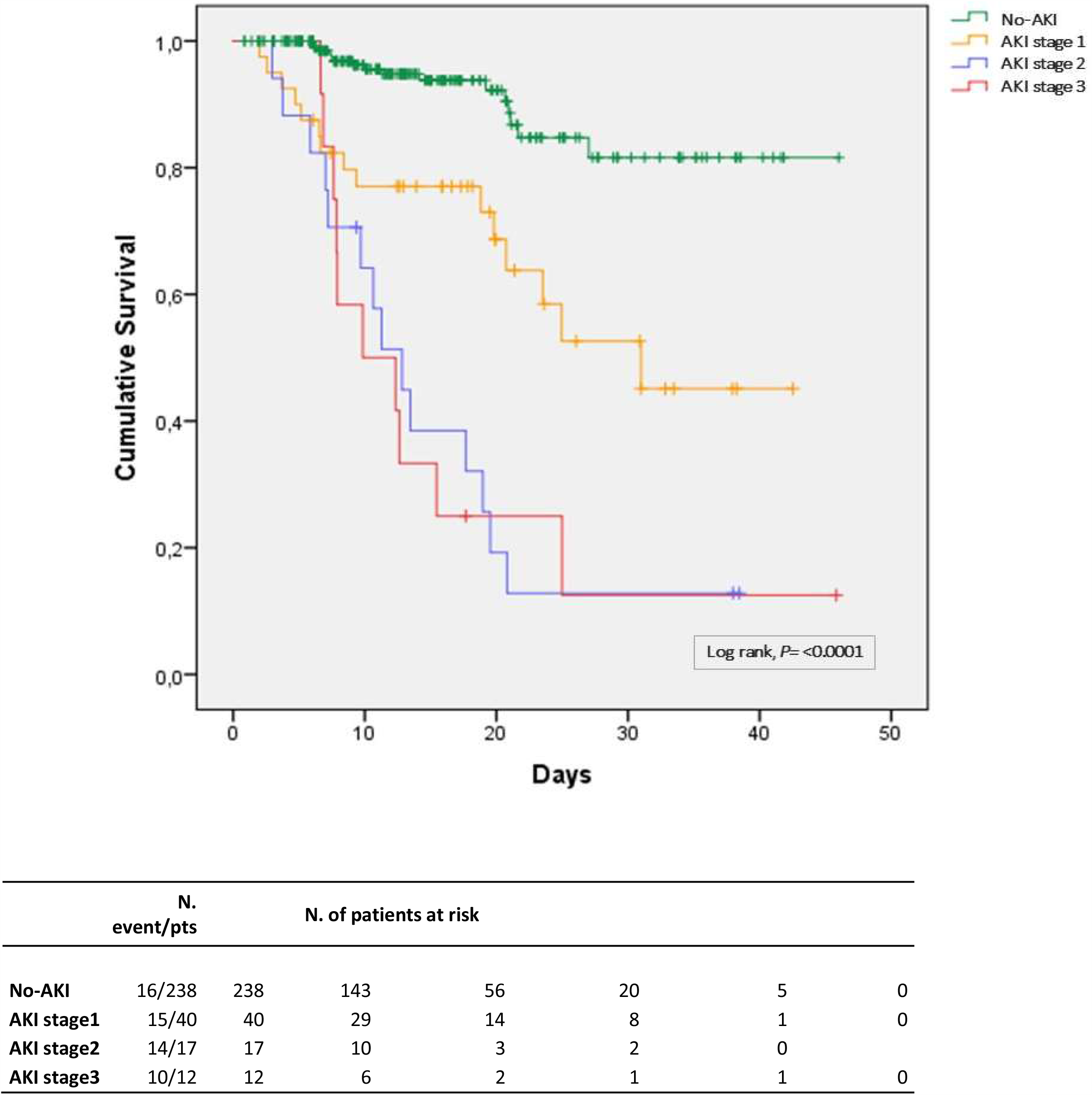
Kaplan Meyer survival analysis between patients with AKI according to KDIGO stage 1-3 and non-AKI.

**Figure 5.**
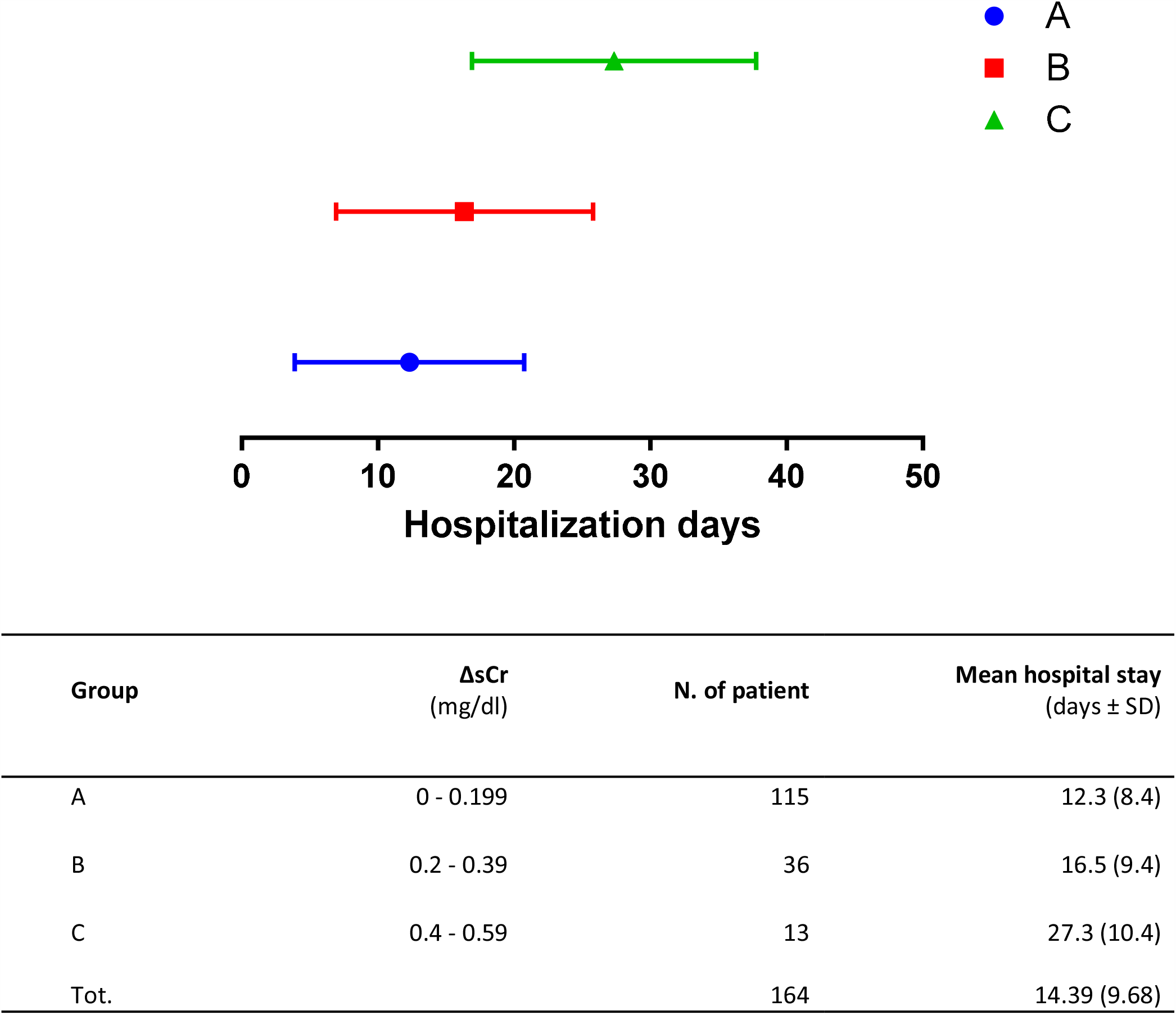
Relationship between the decrease of serum creatinine (sCr) level (mg/dl) and the length of the hospital stay (days). ΔsCr (mg/dl) between baseline and last sCr measurement in non-AKI patients was categorized into A, B and C group. Group A included patients with a ΔsCr ranging from 0 to 0.199; group B included patients with ΔsCr ranging from 0.2 to 0.399 and group C included patients with ΔsCr ranging from 0.4 to 0.599. ANOVA one-way showed a statistically significant difference between the length of hospital stay (days) of the three groups (P= <0.0001). Patients with the highest sCr reduction (group C) had a statistically significant longer hospitalization compared to group A (P=0.0001) and group B (P=0.001).

### Risk factors for AKI

Univariable Cox regression analysis revealed that age (HR=1.064; CI95%, 1.04-1.08), CKD (HR=2.88; CI95%, 1.76-4.71) and non-renal sequential organ failure assessment (SOFA) (HR=1.61; CI95%1.36-1.92) were statistically significant predictors of AKI. Age over 65 years (HR=4.24; CI95%2.23-8.09) and non-renal SOFA > 3 points (HR=5.10; CI%2.41-10.8) were the strongest predictors of kidney injury (Table 4). Multivariable analyses showed that non-renal SOFA (HR=2.52; CI95%,1.21-5.23), age (HR=1.05; CI95%, 1.02-1.08), CKD (HR=1.97; CI95%, 1.10-3.54) and male sex (HR=1.33; CI95%, 1.14-1.56) were independent risk factors for AKI in our patients (Table 5)

**Table 4.**
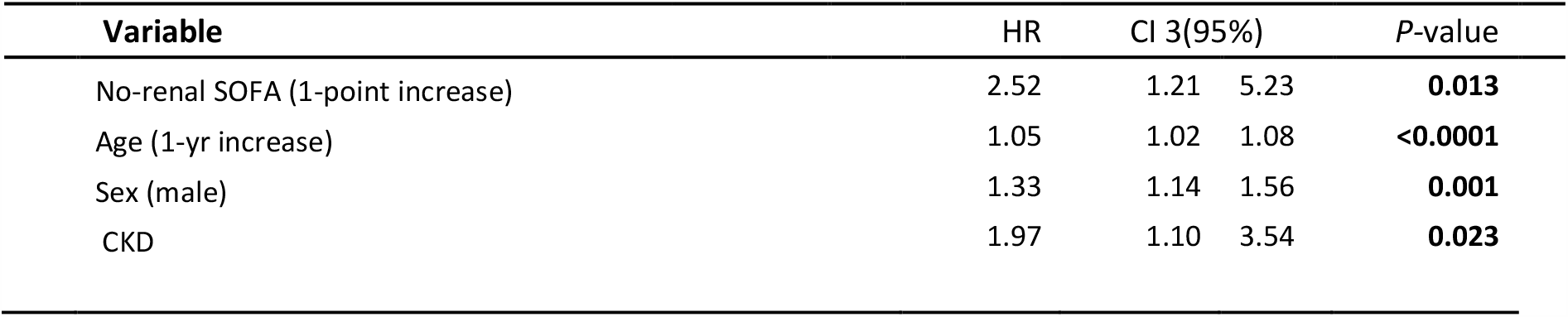
Multivariate Cox Regression analysis to assess risk factors for AKI.

| Variable | HR | CI 3(95%) |  | P-value |
| --- | --- | --- | --- | --- |
| No-renal SOFA (1-point increase) | 2.52 | 1.21 | 5.23 | **0.013** |
| Age (1-yr increase) | 1.05 | 1.02 | 1.08 | **<0.0001** |
| Sex (male) | 1.33 | 1.14 | 1.56 | **0.001** |
| CKD | 1.97 | 1.10 | 3.54 | **0.023** |

**Table 5.** Adjusted and unadjusted Cox proportional hazard regression model for death with 95% confidence intervals between groups of patients

|  | Unadjusted relative hazards of death |  |  |  | Adjusted relative hazards of death* |  |  |  | Adjusted relative hazards of death# |  |  |  |
| --- | --- | --- | --- | --- | --- | --- | --- | --- | --- | --- | --- | --- |
|  | HR | 95% CI |  | *p* value | HR | 95% CI |  | *p* value | HR | 95% CI |  | *p* value |
| No-AKI | 1.00 |  |  |  | 1.00 |  |  |  | 1.00 |  |  |  |
| AKI | 7.184 | 4.010 | 12.869 | <0.0001 | 5.230 | 2.841 | 9.630 | <0.0001 | 3.743 | 1.340 | 10.461 | 0.011 |
|  | HR | 95% CI |  | *p* value | HR | 95% CI |  | *p* value | HR | 95% CI |  | *p* value |
| No-AKI | 1.00 |  |  |  | 1.00 |  |  |  | 1.00 |  |  |  |
| AKI stage 1 | 4.216 | 2.072 | 8.578 | <0.001 | 2.698 | 1.305 | 5.578 | 0,007 | 1.865 | 0.472 | 7.369 | 0.374 |
| AKI stage 2 | 12.944 | 6.302 | 26.589 | <0.0001 | 9.814 | 4.710 | 20.449 | <0.0001 | 1.000 | 0.197 | 5.084 | 0.99 |
| AKI stage 3 | 12.684 | 5.739 | 28.032 | <0.0001 | 11.870 | 4.405 | 31.981 | .00402 | 2.089 | 0.196 | 22.282 | 0.541 |
| AKI stage 2-3 | 12.948 | 6.870 | 24.401 | <0.0001 | 10.323 | 5.307 | 20.080 | <0.0001 | 7.840 | 2.290 | 26.849 | 0.001 |
|  | HR | 95% CI |  | *p* value | HR | 95% CI |  | *p* value | HR | 95% CI |  | *p* value |
| No-AKI | 1.00 |  |  |  | 1.00 |  |  |  | 1.00 |  |  |  |
| Unrecovered AKI | 12.565 | 6.948 | 22.724 | <0.0001 | 10.410 | 5.354 | 20.241 | <0.0001 | 5.378 | 1.754 | 16.490 | 0.003 |

Ci denotes confidential interval
\* Hazard ratio adjusted for age and sex

### Outcome

The primary causes of death were respiratory failure (61.5%), followed by sepsis (15.3%) and septic shock with MOF (9%). Crude mortality was significantly higher in AKI patients (56 vs 6.7%; P=<0.0001) compared to patients with normal renal function (Table 2). KM curves showed high mortality for all patients with AKI, with a significantly worse outcome for patients in AKI stage 2 and 3 (log-rank p<0.0001) (Fig.4) Patients experiencing AKI had an unadjusted HR for death of 7.184 (CI95%4.01-12.86). Adjusted HR for death in the AKI group was 4.2 (CI95%, 2.07-8.57), 12.9 (CI95%, 6.3-26.5) and 12.6 (CI95%, 5.7-28.03) for patients with AKI stages 1, 2, and 3, respectively. In a multivariable COX regression analysis that included age, sex, comorbidities (diabetes mellitus, cardiovascular disease [CVD], chronic kidney disease [CKD], chronic obstructive pulmonary disease [COPD] and non-renal SOFA, the HR for in-hospital death in patients with AKI was 3.74 (CI95%, 1.34-10.46) compared to non-AKI patients.

The risk of in-hospital death was evaluated among the subset of the population with AKI injury. AKI was a significant predictor of all-cause mortality in the group of patients with AKI stage 2 and 3 (HR=7.84; CI95%, 2.29-26.849) and in patients with unrecovered renal function (HR=5.378; CI95%, 1.75-16.49) at the end of the follow-up compared to non-AKI (Table 5).

### Trend of serum creatinine level during hospitalization in non-AKI patients

A positive relationship was identified between the decrease of sCr level and long-term hospitalization (P=<0,0001; r=0,45) in non AKI patients(Supplementary Fig. 1). Indeed, days of hospitalization were a significant predictor of the decrease of sCr (P=<0.0001; decrease of sCr=0.055+0.006 day of hospitalization). Patients with the highest decrease of last sCr respect to baseline (Δ sCr, 0,4-0,599) had a statistically significant longer hospitalization compared to patients of group A (Δ sCr, 0-0,199 mg/dl) and B (Δ sCr, 0,2-0,399) who had a lessen decreases of last sCr value (Fig.5)

## Discussion

The results of this study confirm the recently published data reporting AKI as a frequent event in COVID-19. In a cohort of 307 patients hospitalized for severe respiratory symptoms due to SARS-CoV-2 infection, AKI complicated the clinical course of 22.4% patients. In the majority of patients (57.9%) AKI was mild (stage 1), whereas AKI stage 2 and 3 accounted for 24.6 and 17.3% of the cases, respectively. As already noted in previous studies^10,29^ kidney injury was accompanied by a higher burden of urinary abnormalities such as microscopic hematuria (P=0.032) and proteinuria (0.448 vs 0.18. P=< 0.0001) compared to patients with no renal injury. Renal function was replaced in five subjects with AKI stage 3 by continuous renal replacement therapy. The outcome of these patients was poor because all died without recovering the renal function as a consequence of refractory septic shock evolving in untreatable multiorgan failure. Of note, a worrisome failure to recover from renal injury was noted in over half of the patients (65.2%) at the end of follow-up. It is known that AKI is a devastating syndrome with a significant impact on morbidity and mortality^30^. Early reports from Chinese cohorts documented a low prevalence of renal involvement ^11,31^. Subsequent observational studies conducted in larger cohorts reported an incidence of AKI ranging from 0.5 to 10.4%^11–16^. A recent study evaluating 5449 patients in the New York metropolitan area confirmed that AKI is a frequent complication of acute COVID infection^10^. It was diagnosed in more than one-third of patients with a high burden of comorbidities and, mainly in patients with respiratory distress requiring mechanical ventilation.

We are unable to explain the wide variability between the reported rates, but different criteria adopted for the definition of AKI, population selection, sCr measurement frequency and timing of hospital admission are all potential determinants of these heterogeneous estimates. The close interplay between hemodynamic impairment, inflammation and renal dysfunction^32^ also supports the hypothesis that the severity of the systemic inflammatory host response may play a key role in the development of kidney injury.

In our study, AKI was predominantly diagnosed in symptomatic older male patients (74.7 versus 62.4 years) experiencing a more severe infection compared to no-AKI subjects. Patients who developed AKI presented a significantly higher SOFA score on admission (3.8 versus 2.3), a high level of the classical biomarkers of systemic inflammation (IL-6, LDH, D-Dimer, albumin, platelet count, hemoglobin, ferritin) and impairment of other organs including lung (PO_2_/FiO_2_), heart (troponin, BNP) and liver (bilirubin, ALT).

Of interest, two peaks were noted in the timeline of AKI development. An early peak occurred in the first three days and a second one between days 13 and 15 after admission.

The first peak can be explained by pre-renal causes, as suggested by the New York City (NYC) series^10^. Anorexia, fever and increased perspiration may be all causes of reduced renal perfusion in patients susceptible to acute renal injury. The second AKI peak, roughly coinciding with the death of the patients, may be linked to a critical interplay between deranged microvascular renal perfusion and intrarenal-inflammation due to the pro-inflammatory effect of cytokines in septic patients suffering from hemodynamic collapse. Acute tubular necrosis may be plausible pathogenesis consistent with the severity of the presentation and the late recovery of renal function^18^.

AKI patients carried a higher burden of cardiovascular risk factors compared to the no-AKI group. The incidence of AKI was more frequent among patients with CKD and diabetes mellitus largely known to be associated with an increased vulnerability to kidney injury^33–35^. In the present study, non-renal SOFA, age, male sex and CKD were statistically significant predictors of AKI. The identification of these risk factors may elucidate strategies for the prevention of renal injury, as this event is independently associated in-hospital mortality The burden of this association is estimated with a 7 folds excess risk of mortality in patients in AKI stage 2-3 and 5 fold in subjects with unrecovered AKI. Detection of vulnerable patients at risk for AKI, supportive strategy, and avoidance of nephrotoxic agents in patients prone to AKI could improve the prognosis of these patients and probably prevent long-term consequences.

However, careful attention is required in the diagnosis of “recovery” from AKI as long-hospitalization is associated with a progressive reduction of sCr levels which may mimic recovery of renal function. Muscle wasting generally seen within 10 days from hospitalization^36^, pronounced catabolic effects of proinflammatory cytokines^37^ and reduction of the patient’s body weight with long hospital stay (object of ongoing investigation) corroborate the hypothesis of a low SCr as a marker of muscle depletion.

Several limitations of this study should be mentioned, some of which intrinsic to the retrospective nature of the study. A certain number of AKI events may be underdiagnosed because of the unavailability of the sCr at the time of symptoms onset. Hence, it is likely that the patients enrolled in the non-AKI group had a worse prognosis due to renal impairment. Although we have adjusted for potential demographic and clinical confounding, other unrecognized could not be ruled out. We used the non-renal SOFA in order to avoid collinearity between predictor and outcomes. We are confident that the adjustment of our model for this strong clinical variable reinforces the relationship between AKI and in-hospital mortality. Lastly, the lack of data on consequences of renal injury, do not allow to weight the real long-term consequences of AKI in term of morbidity and mortality in a cohort of patients at high risk for CKD.

## Conclusion

Acute renal injury is a frequent complication of COVID-19, as it tends to occur in one-fifth of patients hospitalized for SARS-COV-2 pneumonia. AKI is generally diagnosed in symptomatic aged patients in a context of severe systemic inflammatory response to the ongoing infection. Non-renal SOFA, age, male sex and CKD were risk factors for AKI in our cohort of patients. Identification of the etiological mechanism of AKI, strategy aimed to prioritize the prevention and early identification of AKI are urgently required, as AKI is an independent predictor of all-cause mortality in COVID- 19.

## Data Availability

Data are available upon reasonable request

---

**Supplementary Table 1.** Proportion of patients with AKI stratified according to the requirement for invasive mechanical ventilation.

|  | Invasive<br>mechanical<br>ventilation<br>(n=53) | No invasive<br>mechanical<br>ventilation<br>(n=254) | *P*-value |
| --- | --- | --- | --- |
| AKI |  |  |  |
| Any stage | 18 (33.9) | 51 (20) | **0.045** |
| Stage 1 | 6 (11.3) | 34 (13.3) | 0.820 |
| Stage 2 | 7 (13.2) | 10 (3.9) | **0.014** |
| Stage 3 | 5 (9.4) | 7 (2.7) | **0.039** |
| Required renal replacement therapy | 3 (5.6) | 2 (0.7) | **0.037** |
| Unrecovered AKI | 12 (22.6) | 33 (12.9) | 0.086 |

AKI denotes acute kidney injury

**Supplementary Figure 1.**
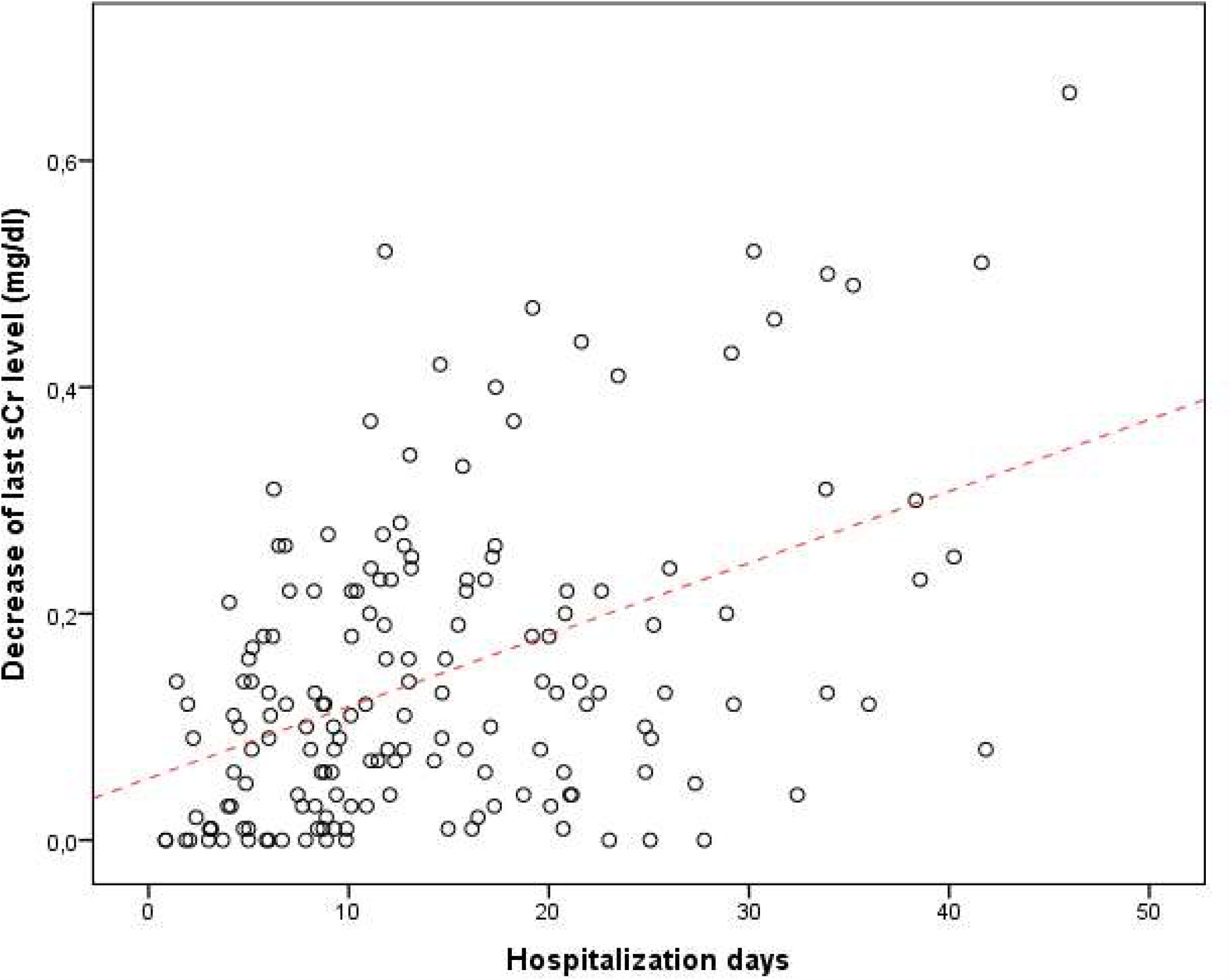
Scatterplots of Δ serum creatinine (sCr) (mg/dl) between last and baseline sCr in the non-AKI group. There is a significant linear positive relationship between reduction of the sCr and hospital stay (P=<0,0001; r=0,4). The increase in the Δ sCr on the y-axis denotes a lower value of the last sCr compared to the baseline sCr. This phenomenon can be linked to muscle wasting due to prolonged hospitalization.

